# Target attainment of extended-interval dosing of tobramycin in patients less than five years of age with cystic fibrosis: A pharmacokinetic analysis

**DOI:** 10.1101/2022.01.02.22268643

**Authors:** Kevin J Downes, Austyn Grim, Laura Shanley, Ronald C Rubenstein, Athena F Zuppa, Marc R. Gastonguay

## Abstract

**Background:** Extended interval dosing (EID) of tobramycin is recommended for treatment of pulmonary exacerbations in adults and older children with cystic fibrosis (CF), but data are limited in patients less than 5 years of age.

**Methods:** We performed a retrospective population pharmacokinetic (PK) analysis of hospitalized children with CF <5 years of age prescribed intravenous tobramycin for a pulmonary exacerbation from March 2011 to September 2018 at our hospital. Children with normal renal function who had ≥1 tobramycin concentration available were included. Nonlinear mixed effects population PK modeling was performed using NONMEM^®^ using data from the first 48 hours of tobramycin treatment. Monte Carlo simulations were implemented to determine the fraction of simulated patients that met published therapeutic targets with regimens of 10-15 mg/kg/day once daily dosing.

**Results:** Fifty-eight patients received 111 tobramycin courses (range 1-9/patient). A 2-compartment model best described the data. Age, glomerular filtration rate, and vancomycin coadministration were significant covariates on tobramycin clearance. The typical values of clearance and central volume of distribution were 0.252 L/hr/kg^0.75 and 0.308 L/kg, respectively. No once daily regimens achieved all pre-specified targets simultaneously in >75% of simulated subjects. A dosage of 13 mg/kg/dose best met the predefined targets of C_max_ >25 mg/L and AUC_24_ of 80-120 mg*h/L.

**Conclusions:** Based on our population PK analysis and simulations, once daily dosing of tobramycin would not achieve all therapeutic goals in young patients with CF. However, extended-interval dosing regimens may attain therapeutic targets in the majority of young patients.

## INTRODUCTION

Pediatric patients with cystic fibrosis (CF) commonly experience pulmonary exacerbations (PEx) for which they receive antimicrobial therapy. Tobramycin, an aminoglycoside, is commonly used as first-line treatment for patients with CF experiencing a PEx. The CF Foundation recommends extended interval dosing (EID) for tobramycin (and other aminoglycosides) to optimize effectiveness (concentration-dependent killing) and reduce the likelihood of safety concerns such as nephrotoxicity.^1^ Although EID has been evaluated, and appears effective and safe in patients 5 years of age and older,^2^ there are limited data to support EID for tobramycin in CF patients less than 5 years of age.

Therapeutic drug monitoring (TDM) can improve both the efficacy and safety of aminoglycosides. These drugs are concentration-dependent antibiotics with post-antibiotic effects,^3^ but are also associated with a variety of dose- and duration-dependent toxicities including nephrotoxicity and ototoxicity. By monitoring aminoglycoside concentrations, providers can adjust doses to ensure that patients receive therapeutic peak serum drug concentrations and appropriate trough concentrations to potentially minimize toxicity.^4^ Providers can also use TDM to determine the drug-free interval (DFI) - the duration of time the drug is undetectable - and ensure that the frequency of drug dosing is suitable. Currently, our institution utilizes once-daily dosing (i.e. EID) of tobramycin for patients with CF 5 years of age and older and every eight-hour dosing for patients less than 5 years of age given the limited data for tobramycin EID in these younger patients with CF.

The optimal initial dose for tobramycin when using EID in young children is unknown. As such, there are concerns about both over- and under-dosing. Younger patients may clear drug more quickly, putting them at a higher risk of inadequate antibiotic therapy when extended dosing intervals are used. Alternatively, large initial doses could result in high initial tobramycin concentrations and result in toxicity. Therefore, informed dosing guidance for tobramycin in children is important. While Arends and colleagues were the first to specifically evaluate EID in a young CF patient population,^5^ their study calculated pharmacokinetic (PK) parameters in individual subjects using noncompartmental methods (i.e. algebraic equations).^5^ This analytic approach using measured drug concentrations can be subject to bias since it does not account for measurement error or inter- or intra-subject variability.

The primary objective of the current study was to utilize a population PK analysis approach to determine the suitability of empiric tobramycin EID in patients less than 5 years of age. Since TDM can be used to guide dosing during therapy, we sought to evaluate early (within 48 hours) PK parameters and assess attainment of CF Foundation-recommended goal tobramycin serum concentrations and DFI through simulations. Secondary objectives of this study were to quantify population PK parameters, including typical values and random inter-individual and residual variabilities, identify important covariates that affect early tobramycin PK, and define drug concentrations attained with EID tobramycin in patients less than 5 years of age with CF. Furthermore, using simulations, we sought to compare target attainment from model predictions to what could be expected in clinical practice using standard TDM approaches.

## METHODS

### Study Design

This was a retrospective, observational study of children with CF less than 5 years of age prescribed intravenous (IV) tobramycin therapy for a PEx. All hospitalized children who received IV tobramycin therapy for standard-of-care treatment between March 1^st^, 2011 and September 1^st^, 2018 at CHOP and had at least one tobramycin concentration measurement performed for TDM were eligible for inclusion. Patients with renal impairment, defined as an estimated glomerular filtration rate (eGFR) <60 mL/min/1.73m^2^ calculated by bedside Schwartz equation,^6^ who received extracorporeal membrane oxygenation (ECMO), with a postmenstrual age of less than 44 weeks, or who received concurrent nebulized tobramycin were excluded. The clinical team determined tobramycin dosing regimens and timing of tobramycin concentration measurements for TDM. The hospital formulary-recommended starting dose for patients less than 5 years of age at CHOP was 3.3 mg/kg/dose IV every 8 hours given as a 30-minute infusion. Peak and trough measurements were routinely obtained following the third dose.

Patients were included into the study for all tobramycin treatment courses received during the study period. Since we were interested in empiric tobramycin dosing for this study, only drug concentrations obtained within the first 48 hours of tobramycin therapy were included. The CHOP Institutional Review Board approved of this study with a waiver of informed consent.

### Data collection

Electronic medical records were reviewed for collection of demographic and biometric characteristics, serum creatinine, concurrent medications, tobramycin dosing information, and tobramycin concentrations. Data were collected in Excel (Microsoft Corp., Redmond, WA). Tobramycin measurements were performed in the CHOP Chemistry Laboratory (CLIA-certified) using competitive immunoassay (VITROS^®^ Chemistry Products TOBRA Reagent, Ortho-Clinical Diagnostics, Inc., Rochester, NY) throughout the study period. The lower limit of quantification of this assay was 0.6 mg/L.

### Data analysis

#### Base model

Population PK analyses were conducted using nonlinear mixed-effects modeling with NONMEM^®^ software v7.4 and the PDx-Pop v5.2.1 interface (ICON plc. Dublin, Ireland). With prior knowledge on the compartmental disposition of tobramycin,^7^ we sought to develop a 2-compartment model with first-order elimination. Since our study relied on TDM peak and trough data, we recognized that the current study design would not support estimation of parameters for the known tobramycin 2-compartment pharmacokinetic model disposition. Given that, a literature review identified published extensively sampled tobramycin PK data that could serve to support estimation of the current pediatric population PK model parameters and covariance terms.^8^ This prior information was quantified by estimating the population PK model parameters for the literature data. The prior knowledge was formally incorporated in the current analysis by utilizing a penalized likelihood function with parametric specification of informative prior distributions on selected parameters from the literature model, and estimation of maximum *a posteriori* probability (MAP) Bayesian population PK parameters given the current TDM data set, as has been described previously.^9^

Between-subject variability was modeled using exponential variability, which assumes log-normal distribution of between subject variability around a parameter. A full block covariance matrix was utilized to define the between-subject random effects. Residual variability (RV) was estimated using a proportional error model. Model selection was driven by the data and based on various goodness of fit indicators, including comparisons based on the minimum objective function value (OFV), visual inspection of diagnostic scatter plots, and evaluation of estimates of population fixed and random effect parameters.

#### Handling of BQL data

Omission of below quantification limit (BQL) data can introduce substantial bias in parameter estimates,^10,11^ particularly when there is a large amount of missing data (>10-15%). To minimize bias associated with BQL data, we developed models using the M3 method described by Beal.^12^

#### Covariate selection

Initially, covariate-parameter relationships were explored graphically and any correlations between covariates noted. Covariate model selection was then conducted by a stepwise backward elimination technique starting with a full covariate model, which was carefully constructed to avoid inclusion of collinear or correlated covariates. Based on previous population PK studies of children with CF treated with tobramycin,^7^ covariates included *a priori* were patient age and eGFR (calculated using Schwartz equation) on CL.^6^ Age was evaluated as a Hill function on CL, as well as an exponential covariate normalized to the population median. No covariates were tested on volume parameters due to the absence of associations in published pediatric models aside from weight.^7^ During backwards elimination, a critical change in OFV of ≥ 6.63 for the FOCE method (nominal α=0.01, df=1) was used to determine covariate inclusion.

Following this initial covariate evaluation process, we then performed an exploratory covariate analyses to assess the effects of concurrent nephrotoxic medication treatment on tobramycin CL. The influence of vancomycin, ticarcillin/clavulanate, trimethoprim/sulfamethoxazole, piperacillin/tazobactam, and NSAID co-administration was evaluated using a forward selection process (a reduction in OFV of ≥ 6.63 for inclusion). Each nephrotoxin exposure was dichotomized (Y/N) as a time-varying covariate, allowing it to vary over the course of tobramycin. We further explored the influence of nephrotoxin exposures as a class effect by grouping agents together as: none vs ≥1 agent, and 0-1 vs ≥2 agents.

#### Inter-occasion variability (IOV)

To account for multiple treatment courses for the same patient over different hospitalizations, we assessed occasion-specific random effects on the structural model parameter of CL. We defined an occasion as a unique hospitalization per subject; if a patient received multiple treatment courses during a single hospitalization, only the first treatment course was included. We compared the impact of inclusion of IOV on model fit (AIC) between the final covariate model with and without IOV, evaluated the influence of IOV on the fixed and random effects parameter estimates, and examined its impact on residual unexplained variability.

#### Model performance

Model selection was determined by evaluating goodness-of-fit diagnostic plots, comparisons of the minimum objective function value (OFV), Akaike information criterion (AIC), and precision of parameter estimates. Visual predictive checks (VPCs) were performed to assess the fit of the final covariate model. 500 VPC simulations were obtained from the observed data. Observed tobramycin data was plotted along with the 95th, 50th and 5^th^ percentiles for the simulated data sets. Data was plotted as time after dose (TAD) vs. concentration. RStudio v3.6.3 (RStudio, Inc., Boston, MA) was used for descriptive statistics and graphical evaluations.

#### Simulations

Monte Carlo simulations were performed to project concentrations achieved with EID dosing of 10-15 mg/kg/day. Simulations were based on relevant study population characteristics (e.g. age, weight, GFR, etc.) at the start of their first tobramycin course and the final PK model estimates. A total of 500 simulations were run for each subject in the study population, generating 29,000 simulated subjects, using two approaches. First, random residual variability was fixed to 0 to allow our simulations to reflect only random between subject PK variability within the population. Second, we included the random residual variability from our population PK model to better reflect variability present within a clinical sample observation context.

#### Target attainment

Using the simulation output, we then determined how often simulated subjects met the following *a priori* goal parameters set forth by the CF Foundation for EID^1^: tobramycin peak (C_max_) concentration of ≥25 mg/L, tobramycin trough (C_min_) concentration of <0.6 mg/L, DFI of <11 hours, and a 24-hour area under the curve (AUC_24_) of 80-120 mg*h/L. Since increased tobramycin exposure is associated with nephrotoxicity, we further determined the proportion of simulated subjects with an AUC_24_ > 120 mg*h/L. Target attainment was assessed following the 2^nd^ EID dose in two ways.

First, using the simulations with zero residual variability, C_max_, C_min_, and DFI targets were determined directly from the simulated concentrations, while AUC_24_ was calculated using the equation, AUC_24_ = daily dose/CL, using each simulated subject’s dose and CL estimates. This equation assumes drug is at steady-state, which we felt was a reasonable assumption for patients receiving EID. This approach is called the “Simulation Output” approach in results below. Second, using the simulations that incorporated residual variability from our population PK model, we performed log-linear regression on simulated concentrations at 3 and 8 hours to calculate AUC_24_ C_max_, C_min_, and DFI targets. The equations used for this approach are shown in the Supplemental Materials. The log-linear approach is used clinically for TDM at many institutions,^13^ including ours, so this approach was called the “TDM Approach.” The results of these two methods demonstrate differences between model expectations and clinical calculations in a real-world setting.

## RESULTS

### Study population

Sixty-one patients received 115 tobramycin courses during the study period. Four courses in 3 patients were excluded (two in premature infants, two with no tobramycin concentrations obtained). Thus, 58 patients receiving 111 tobramycin courses were included. **Table 1** displays baseline characteristics of the study population during their first course and all courses during the study. The median age of patients at the initiation of therapy was 2.2 years (IQR: 1.2-4). Thirty-five subjects received one tobramycin course during the study period, 13 received two courses, and 10 received three or more courses. Two-hundred twenty-eight tobramycin serum concentrations were collected; 4 concentrations were excluded due to the samples being mistimed. Of 224 tobramycin concentrations included, 53 (23.7%) were reported as BQL. **Figure 1** depicts the distribution of tobramycin serum concentrations in relation to time after dose.

**Table 1.** Characteristics of study population based on first course and all courses of tobramycin.

| Characteristic | First course (n=58) | All courses (n=111)<sup>a</sup> |
| --- | --- | --- |
| Female gender, n (%) | 25 (43%) | 46 (41%) |
| Age in years, median (IQR) | 2.2 (0.8 - 3.8) | 2.6 (1.2 - 4) |
| Weight in kg, median (IQR) | 11.2 (8.2 – 14.7) | 12.4 (9.7 – 14.8) |
| Height in cm, median (IQR) | 85.8 (70.5 – 95.5) | 88.1 (75 – 98) |
| Serum creatinine in mg/dL, median (IQR)<sup>b</sup> | 0.3 (0.2 – 0.3) | 0.3 (0.2 – 0.3) |
| GFR in ml/min/1.73m<sup>2</sup>, median (IQR)<sup>b</sup> | 126 (110– 149) | 129 (110 – 148) |
| GFR < 90 ml/min/1.73m<sup>2</sup>, n (%)<sup>b</sup> | 3 (5.2%) | 6 (5.4%) |
| Dose in mg/kg/dose, median (IQR)<sup>b</sup> | 3.2 (3.1 – 3.3) | 3.2 (3.1 – 3.3) |
| Concurrent nephrotoxic medications, n (%)<sup>b,c</sup> |  |  |
| 0 | 38 (65.5) | 70 (63.0) |
| 1 | 18 (31.0) | 33 (29.7) |
| 2 or more | 2 (3.4) | 8 (7.2) |

<sup>a</sup> Among 58 individuals treated, 35 received 1 course, 13 received 2 courses, 4 received 3 courses, 1 received 4 courses, 2 received 5 courses, and 1 each received 7, 8 and 9 courses.
<sup>b</sup> At time of tobramycin initiation
<sup>c</sup> Concurrent nephrotoxic medications included non-steroidal anti-inflammatory medications, acyclovir, vancomycin, piperacillin/tazobactam, ticarcillin/clavulanate, and trimethoprim/sulfamethoxazole.

**Figure 1:**
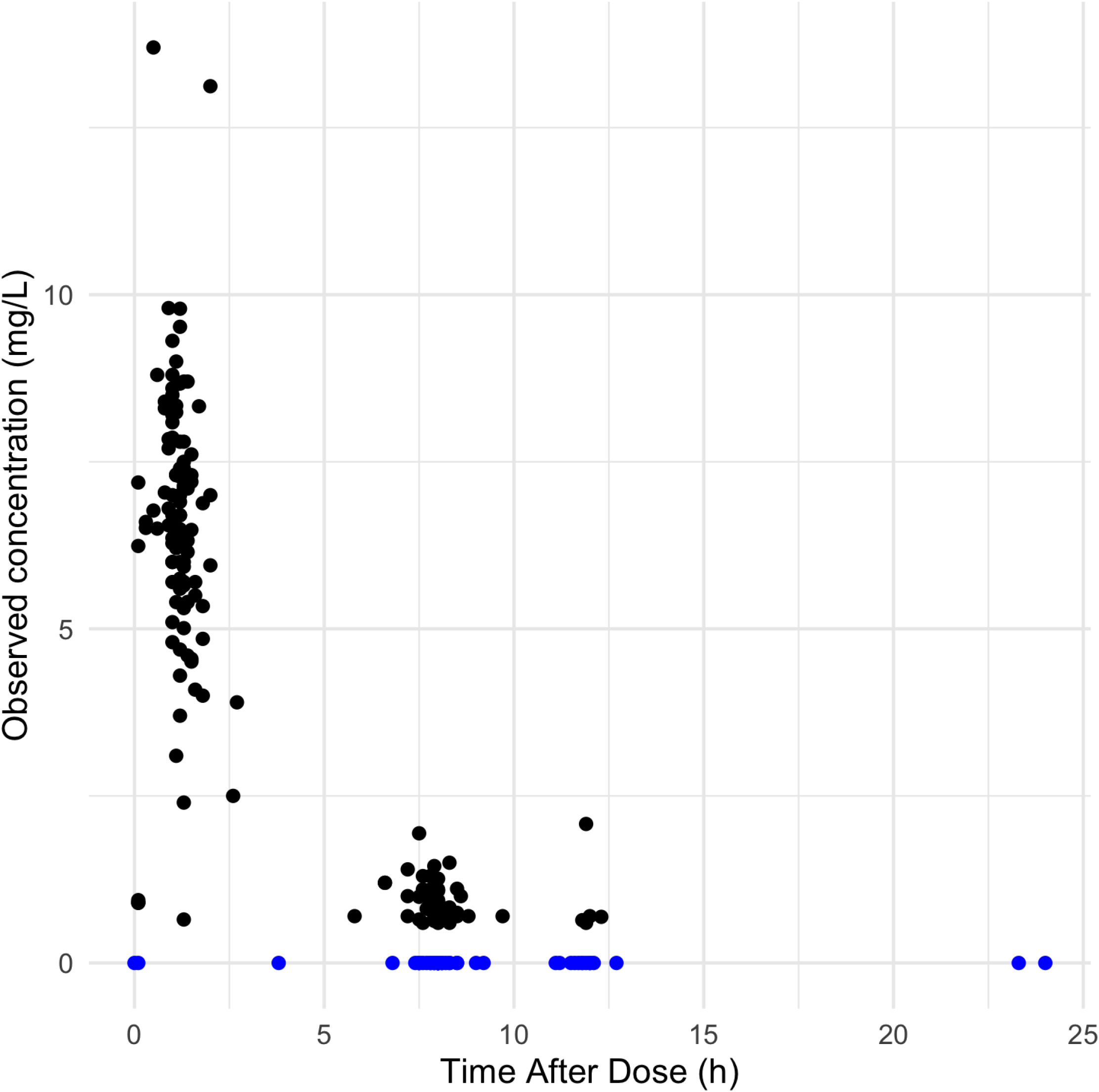
Distribution of tobramycin measurements in relation to time after dose. Drug concentrations reported as below limit of quantification are shown in blue and plotted as 0 mg/L.

### Model development

Due to the timing of TDM sampling in our dataset (i.e. peaks and troughs), we utilized prior information from adult CF patients to inform initial parameter estimates for Q and V2. We identified a published, standard two-stage PK analysis that included rich sampling in 6 adult CF patients treated with 3.3 mg/kg thrice daily tobramycin.^8^ Because all model parameters were not reported in this publication, we used WebPlotDigitizer^14^ to extract the concentration-time data of each subject in Figure 2 of this published model.^8^ We then constructed a dataset for analysis in NONMEM, using median population weight for each subject, to derive initial Q and V2 estimates for our model. Given poor estimation of between-subject random effects for Q and V2 during base model development, they were fixed to 0 for these parameters.

**Figure 2:**
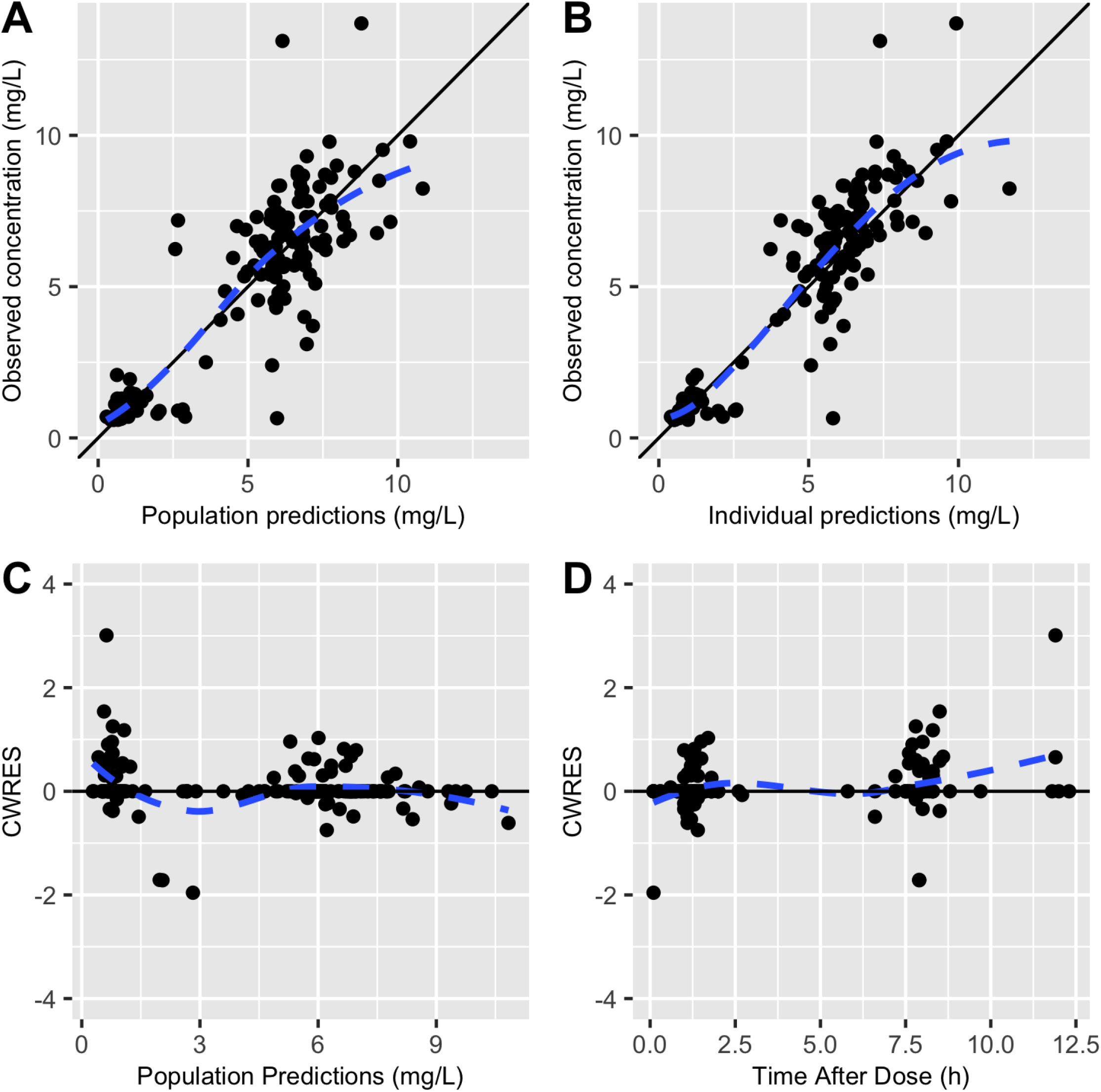
Diagnostic plots for final population pharmacokinetic model. A: Observed vs. population-predicted concentrations. B: Observed vs. individual predicted concentrations. C. Conditional weighted residuals (CWRES) vs. population-predicted concentrations. D. CWRES vs time after dose. Observed concentrations below limit of quantification omitted.

Clearance (CL) and inter-compartmental clearance (Q) were allometrically scaled for weight to 0.75, normalized to 70-kg, while central (V1) and peripheral (V2) volume parameters were scaled linearly for weight, normalized to 70-kg.^15^ The results of the covariate selection process are shown in **Supplemental Table 1**. Age and eGFR were included in the model *a priori*, based on prior published models.^7^ Concomitant receipt of vancomycin was also informative on CL in the forward stepwise approach. When evaluating IOV, the addition of IOV increased the AIC of the model, had no impact on inter-individual random effects on CL and V, and did not change the point estimates of the model parameters. Additionally, IOV minimally reduced the residual unexplained variability of the model from 33.4% to 32.8% and led to an increase in the percent relative standard error for each parameter estimate, suggesting that inclusion of IOV did not improve model fit. Therefore, IOV was not included in our final model.

The parameter estimates for the final model are shown in **Table 2**, along with bootstrap estimates (n=1000 replications) of parameters with 95% confidence intervals. Additional bootstrap analyses were performed (n=500) with stratification by receipt of vancomycin, by age (<1 year, 2-3 years, or 4+ years), and by eGFR (<100, 100-199, 200+ mL/min/m^2^). In each case, the model parameter estimates were within the 95% confidence intervals of the bootstrap estimates. The associated diagnostic plots for the final model are shown in **Figure 2**.

**Table 2.**
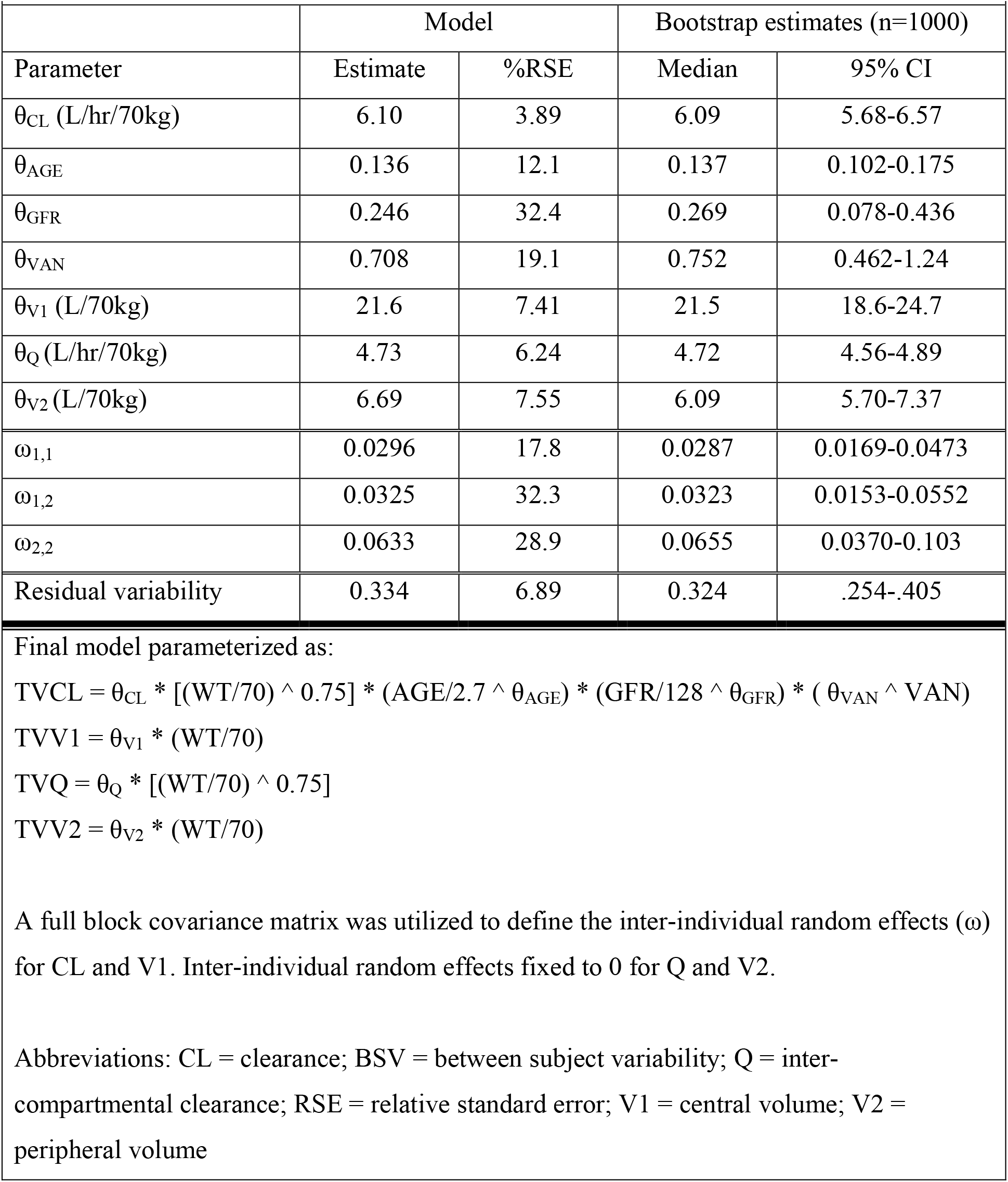
Final population PK parameter estimates.

In the final model, the typical value of CL for our population was 0.252 L/hr/kg^0.75 (95% CI: 0.233 – 0.271), V1 was 0.308 L/kg (95% CI: 0.264 - 0.353), Q was 0.195 L/hr/kg^0.75 (95% CI: 0.171 – 0.219), and V2 was 0.096 L/kg (95% CI: 0.81 – 0.110). Concomitant receipt of vancomycin was associated with a 29.2% (95% CI: 3.7-55.7%) reduction in tobramycin CL, although vancomycin was administered to only 5 patients during 8 tobramycin courses. **Supplemental Figure 1** displays the VPCs for the final covariate model. The median and 5^th^ and 95^th^ percentiles of the predicted tobramycin concentrations fit closely to the observed data, suggesting a good model fit.

### Target Attainment

Monte Carlo simulations were performed for tobramycin dosing regimens of 10-15 mg/kg once daily. Due to the small number of patients in our original study population who received concomitant vancomycin treatment, all simulated patients did not receive vancomycin. **Table 3** reports the proportion of simulated patients achieving the goal parameters of EID.

**Table 3.**
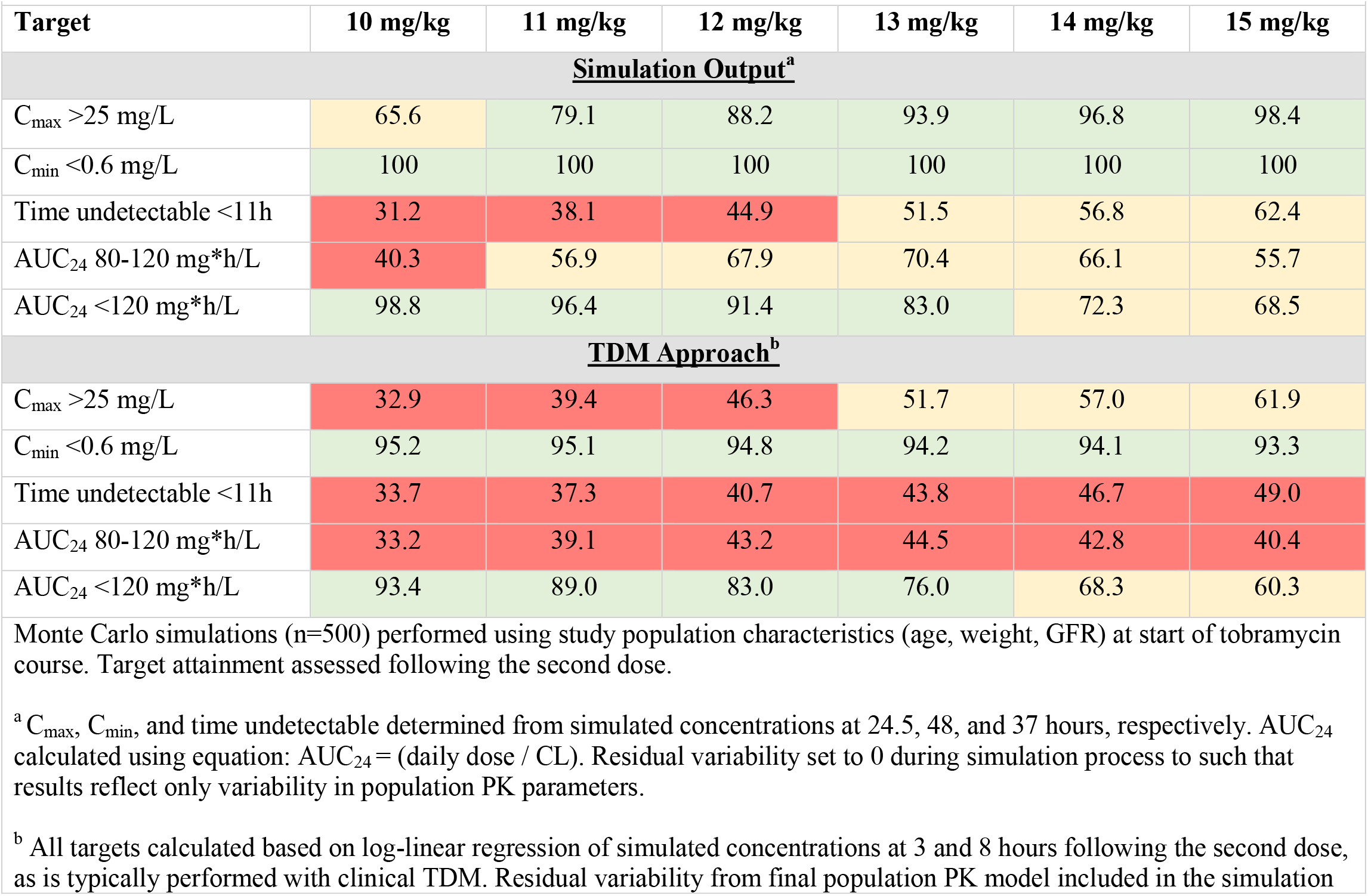
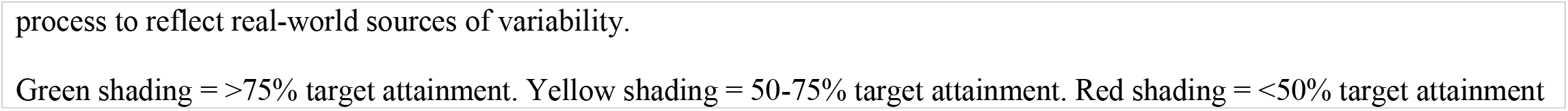
Percentage of simulated patients meeting therapeutic targets on day 2 of treatment based on simulated concentrations.

| Target | 10 mg/kg | 11 mg/kg | 12 mg/kg | 13 mg/kg | 14 mg/kg | 15 mg/kg |
| --- | --- | --- | --- | --- | --- | --- |
| **<u>Simulation Output<sup>a</sup></u>** |  |  |  |  |  |  |
| C<sub>max</sub> >25 mg/L | 65.6 | 79.1 | 88.2 | 93.9 | 96.8 | 98.4 |
| C<sub>min</sub> <0.6 mg/L | 100 | 100 | 100 | 100 | 100 | 100 |
| Time undetectable <11h | 31.2 | 38.1 | 44.9 | 51.5 | 56.8 | 62.4 |
| AUC<sub>24</sub> 80-120 mg*h/L | 40.3 | 56.9 | 67.9 | 70.4 | 66.1 | 55.7 |
| AUC<sub>24</sub> <120 mg*h/L | 98.8 | 96.4 | 91.4 | 83.0 | 72.3 | 68.5 |
| **<u>TDM Approach<sup>b</sup></u>** |  |  |  |  |  |  |
| C<sub>max</sub> >25 mg/L | 32.9 | 39.4 | 46.3 | 51.7 | 57.0 | 61.9 |
| C<sub>min</sub> <0.6 mg/L | 95.2 | 95.1 | 94.8 | 94.2 | 94.1 | 93.3 |
| Time undetectable <11h | 33.7 | 37.3 | 40.7 | 43.8 | 46.7 | 49.0 |
| AUC<sub>24</sub> 80-120 mg*h/L | 33.2 | 39.1 | 43.2 | 44.5 | 42.8 | 40.4 |
| AUC<sub>24</sub> <120 mg*h/L | 93.4 | 89.0 | 83.0 | 76.0 | 68.3 | 60.3 |

Monte Carlo simulations (n=500) performed using study population characteristics (age, weight, GFR) at start of tobramycin course. Target attainment assessed following the second dose.
<sup>a</sup> C<sub>max</sub>, C<sub>min</sub>, and time undetectable determined from simulated concentrations at 24.5, 48, and 37 hours, respectively. AUC<sub>24</sub> calculated using equation: AUC<sub>24</sub> = (daily dose / CL). Residual variability set to 0 during simulation process to such that results reflect only variability in population PK parameters.
<sup>b</sup> All targets calculated based on log-linear regression of simulated concentrations at 3 and 8 hours following the second dose, as is typically performed with clinical TDM. Residual variability from final population PK model included in the simulation
process to reflect real-world sources of variability.
Green shading = >75% target attainment. Yellow shading = 50-75% target attainment. Red shading = <50% target attainment

When assessing attainment of targets determined directly from the Simulation Output approach (i.e. using simulated concentrations and clearance estimates; no residual variability), all simulated patients achieved the C_min_ goal of <0.6 mg/L. And, more than 75% of simulated patients achieved the C_max_ goal of >25 mg/L with 11-15 mg/kg/day dosing regimens. No EID regimens achieved the AUC target (80-120 mg•h/L) in >75% of simulated patients. More than 75% of simulated patients had an AUC >80 mg•h/L with dosing of 12-15 mg/kg/day, although an increasing proportion of simulated subjects’ AUC exceeded 120 mg•h/L at higher dosages. Across all 5 targets, a dosage of 13 mg/kg had the highest average target attainment (79.8%), followed by 12 mg/kg (78.5%) and 14 mg/kg (78.4%).

There were substantial differences in target attainment when the TDM Approach was assessed (i.e. log-linear regression on simulated concentrations at 3- and 8-hours; incorporated residual variability). In general, all targets were less often met using the log-linear approach compared to model expectations. The differences were most significant for C_max_ and AUC targets. At a dose of 13 mg/kg, 42% fewer simulated subjects met the C_max_ target using the log-linear estimation approach compared to the simulation output, while 26% fewer met the AUC_24_ target of 80-120 mg*h/L.

## DISCUSSION

This retrospective, population PK study analyzed the applicability of EID for IV tobramycin in CF patients less than five years of age, and compared attainment of therapeutic targets in simulated patients according to CF Foundation guidelines.^1^ Our patient cohort included a wide variety of CF patients in terms of age and prior tobramycin exposures. Our simulations suggest that EID tobramycin would not attain all recommended therapeutic goals in a significant majority (>75%) of patients with CF less than five years of age. However, a dosage of 13 mg/kg/day had the highest probability of target attainment across all 5 targets evaluated and would be the optimal EID dosing regimen, according to our simulations. This dose met the C_max_ and C_min_ targets in >90% of simulated subjects, while also achieving the time undetectable and AUC_24_ targets in >50% and >70%, respectively.

EID can maximize the concentration-dependent effectiveness of tobramycin, while minimizing its risk of renal toxicity associated with accumulation of drug within proximal tubule cells in setting of sustained exposure.^16^ EID tobramycin in patients older than five years of age has comparable efficacy and improved safety compared to conventional dosing in older children,^2,17,18^ but data in younger children are limited. Arends and colleagues retrospectively evaluated tobramycin EID in 31 CF patients less than 6 years of age,^5^ and found that eight out of the 29 patients evaluated for nephrotoxicity developed kidney injury according to pediatric RIFLE definitions;^19^ this is a higher rate than reported in other studies of acute kidney injury (AKI) in older children with CF treated with tobramycin,^20^ although only 2 patients had increases in serum creatinine of ≥0.2 mg/dL,^5^ which could be considered a clinically significant change. Although these authors concluded that dosing of tobramycin at 12 mg/kg/dose once daily would achieve recommended peak concentrations, the effectiveness of this EID has not been fully evaluated in young children and remains an important question. Based on our simulations, a dosage of 13 mg/kg/dose appears optimal, but prospective studies are needed.

We focused on tobramycin PK in the first two days of treatment to determine if EID would be appropriate for empiric use in young children with CF. In older children who receive EID tobramycin at our institution, TDM is routinely performed following the first or second dose, and then weekly during the course of treatment. Our analysis mirrored this approach by examining early PK and assessing target attainment on day 2. There was a substantial degree of variability between the patients within our study population. As a result, EID regimens with higher dosages obtained C_max_ and C_min_ concentration targets in greater than 75% of simulated patients. However, similar results were not found for the percent of patients achieving DFI and AUC goals. With extended durations of DFIs, patients may have prolonged periods of low tobramycin concentrations that put them at theoretical risk of antimicrobial failure. A large proportion of the simulated patients at higher dosages (14-15 mg/kg/day) who did not meet the AUC target did so due to supra-therapeutic AUC values (>120 mg*h/L), which could result in elevated risks of tobramycin toxicity.

Importantly, there were substantial differences in target attainment depending on the method used. These were most notable for estimates of C_max_ and AUC_24_, the two parameters that describe tobramycin exposure. In the absence of Bayesian methods, log-linear regression is often used for clinical TDM to estimate PK parameters and target attainment in individual patients.^13^ Since this approach relies on assumptions of a one-compartment model with linear elimination, C_max_ can be grossly underestimated for a two-compartment drug, as we found. The log-linear approach can also lead to significant differences in AUC_24_ estimates compared to Bayesian methods. In a retrospective study of 77 children with CF treated with once daily tobramycin at a mean dose of 12.5 mg/kg/day, Brockmeyer et al. found that 52% met the AUC_24_ target of 80-120 mg*h/L after the initial dose based on log-linear regression.^13^ This is comparable to our analyses, in which 43-44% of simulated patients given 12-13 mg/kg/day met this AUC target with log-linear regression. Brockmeyer also utilized Bayesian forecasting, based on a two-compartment model, to estimate AUC_24_ for each subject.^13^ Again, similar to our assessment, estimates of AUC_24_ using Bayesian methods were higher than with log-linear regression (mean difference of 6.4 mg*h/L).^13^ These findings have significant clinical ramifications. Dose adjustments based on log-linear regression estimates of C_max_ and AUC_24_ may lead to over-exposure and added risk of toxicity in some patients. Clinicians therefore need to be cognizant of the limitations of traditional TDM practices.

Our estimates for CL and central volume are similar to previous studies involving children with CF treated with EID tobramycin. Massie et al. studied 44 children aged 9 months to 20 years given 12 mg/kg/day.^21^ Based on a one-compartment model, they found that CL and volume of distribution were 0.103 L/h/kg and 0.267 L/kg, respectively.^21^ Hennig et al. performed a population PK analysis of 35 patients with CF aged 0.5 to 17.8 years treated with 10 mg/kg/day of tobramyin.^22^ Their two-compartment model estimated median population values for clearance of 6.37 L/h/70 kg and central volume of 18.7 L/70 kg,^22^ again similar to our model’s estimates (Table 2). However, in a study involving 85 children with CF 5-15 years of age by Touw et al.,^23^ volume of distribution was larger among recipients of once daily compared to thrice daily therapy (0.401 vs 0.354 L/kg, respectively). Clearance was not reported, but elimination rate did not differ by dosing regimen in this study.^23^ So, while the findings of our study are consistent with prior reports,^21,22^ it is possible that PK parameters could have differed in our population of young children had they actually been administered EID.

Because TDM concentrations were collected as standard of care, approximately 28% of the tobramycin samples included in our study were reported as BQL. While we attempted to account for this using the validated M3 method described by Beal,^12^ this may have contributed to increased variability in PK parameters in the model. Omission of BQL data can introduce substantial bias in parameter estimates. In our study, omission of BQL would result in under-estimation of CL since the vast majority of BQL data were trough measurements. The M3 method estimates the likelihood that BQL data are actually below quantification limits and is associated with less bias than omission of BQL data when missing data are during the elimination phase. Thus, despite a substantial amount of BQL data in our study, we believe that our PK parameter estimates are accurate.

Since this study was retrospective and relied on standard-of-care TDM drug concentrations, multiple additional limitations exist, which also may have contributed to variability in the model estimates. First, documentation of the duration of infusion was not a component of standard institutional practice during the study period. However, CHOP utilized standard infusion procedures, including a routine infusion time (30 minutes), which was assumed to have been used in all cases in PK modeling. Incorrect recording of the timing of tobramycin sampling or administration similarly could have affected the estimated PK parameters in the model. Any tobramycin samples that were deemed to clearly be an error (i.e. concentrations drawn during an infusion, concentrations reported as undetectable within 2 hours after the infusion) were not included in the analysis. Lastly, the number of tobramycin serum samples varied greatly between patients, which could have led to differential patient contribution in the PK model.

Overall, EID regimens optimize tobramycin efficacy while potentially lessening toxicity risk in subjects five years and older. Based on our population PK analysis and simulations, EID regimens may not achieve all recommended pharmacokinetic targets for younger children with CF. However, certain doses of EID can optimize the effectiveness and safety of tobramycin in patients less than five years of age while achieving targets in most individuals. Use of TDM will be paramount to ensure that appropriate dosing regimens are administered during prolonged courses, but clinicians should recognize that traditional TDM practices using log-linear regression methods underestimate the true C_max_ and AUC_24_ in patients. Prospective studies will be needed to validate our findings and specifically evaluate the safety and effectiveness of these regimens in CF patients less than five years of age.

## Data Availability

All data produced in the present study are available upon reasonable request to the authors

## Abbreviations

AIC: Akaike information criterion
AUC: area under the concentration-time curve
BQL: below quantification limit
CF: cystic fibrosis
CHOP: the Children’s Hospital of Philadelphia
CL: clearance
C_max_: maximal concentration
C_min_: minimum concentration
DFI: drug-free interval
EID: extended interval dosing
IOV: inter-occasion variability
OFV: objective function value
PD: pharmacodynamic(s)
PK: pharmacokinetic(s)
Q: inter- compartmental clearance
TDM: therapeutic drug monitoring
VPC: visual predictive check
V1: central volume
V2: peripheral volume

## FUNDING

This study was carried out as part of our routine work. KJD is supported by the Eunice Kennedy Shriver National Institute of Child Health & Human Development of the National Institutes of Health under Award Number K23HD091365.

## TRANSPARENCY DECLARATIONS

KJD has received research support from Merck & Co., Inc. unrelated to the current work. AFZ has received research support from Eunice Kennedy Shriver National Institute of Child Health & Human Development (award numbers UG1HD063108, R21HD093369), the U.S. Department of Defense (award number W81XWH-17-1-0668), and Zelda Therapeutics, unrelated to this work. The content is solely the responsibility of the authors and does not necessarily represent the official views of any of the above supporting agencies. All other authors: Nothing to declare.

## Figure Legends

**Supplemental Figure 1: Visual predictive check of concentrations versus time after dose**. Observed tobramycin time after dose compared with the 95th, 50th and 5^th^ percentiles for 500 simulated data sets. Comparison of median (solid blue line) and 5th–95^th^ quantiles (red dashed lines). The 50^th^ percentile prediction interval is depicted as the solid blue band and the 5^th^ and 95^th^ percentile prediction intervals are depicted as red bands. Observed values below limit of quantification were omitted. Simulated values <0.6 mcg/mL refit at 0.6 mcg/mL since observed values cannot be quantified below this threshold.

## Notes

### Competing Interest Statement

All potential COI has been disclosed in the manuscript.

### Funding Statement

This study did not receive any funding

### Author Declarations

IRB of the Childrens Hospital of Philadelphia gave ethical approval for this work.

## REFERENCES

1. Flume PA, Mogayzel PJ, Jr., Robinson KA, et al. Cystic fibrosis pulmonary guidelines: treatment of pulmonary exacerbations. Am J Respir Crit Care Med. 2009;180(9):802–808.

2. Smyth A, Tan KH, Hyman-Taylor P, et al. Once versus three-times daily regimens of tobramycin treatment for pulmonary exacerbations of cystic fibrosis--the TOPIC study: a randomised controlled trial. Lancet. 2005;365(9459):573–578.

3. Huth ME, Ricci AJ, Cheng AG. Mechanisms of aminoglycoside ototoxicity and targets of hair cell protection. Int J Otolaryngol. 2011;2011:937861.

4. Roberts JA, Norris R, Paterson DL, Martin JH. Therapeutic drug monitoring of antimicrobials. Br J Clin Pharmacol. 2012;73(1):27–36.

5. Arends A, Pettit R. Safety of Extended Interval Tobramycin in Cystic Fibrosis Patients Less an 6 Years Old. J Pediatr Pharmacol Ther. 2018;23(2):152–158.

6. Schwartz GJ, Work DF. Measurement and estimation of GFR in children and adolescents. Clin J Am Soc Nephrol. 2009;4(11):1832–1843.

7. Bloomfield C, Staatz CE, Unwin S, Hennig S. Assessing Predictive Performance of Published Population Pharmacokinetic Models of Intravenous Tobramycin in Pediatric Patients. Antimicrob Agents Chemother. 2016;60(6):3407–3414.

8. Aminimanizani A, Beringer PM, Kang J, Tsang L, Jelliffe RW, Shapiro BJ. Distribution and elimination of tobramycin administered in single or multiple daily doses in adult patients with cystic fibrosis. J Antimicrob Chemother. 2002;50(4):553–559.

9. Knebel W, Gastonguay MR, Malhotra B, El-Tahtawy A, Jen F, Gandelman K. Population pharmacokinetics of atorvastatin and its active metabolites in children and adolescents with heterozygous familial hypercholesterolemia: selective use of informative prior distributions from adults. J Clin Pharmacol. 2013;53(5):505–516.

10. Bergstrand M, Karlsson MO. Handling data below the limit of quantification in mixed effect models. AAPS J. 2009;11(2):371–380.

11. Gastonguay MR, French JL, Heitjan DF, Rogers JA, Ahn JE, Ravva P. Missing data in model-based pharmacometric applications: points to consider. J Clin Pharmacol. 2010;50(9 Suppl):63S-74S.

12. Beal SL. Ways to fit a PK model with some data below the quantification limit. J Pharmacokinet Pharmacodyn. 2001;28(5):481–504.

13. Brockmeyer JM, Wise RT, Burgener EB, Milla C, Frymoyer A. Area under the curve achievement of once daily tobramycin in children with cystic fibrosis during clinical care. Pediatr Pulmonol. 2020;55(12):3343–3350.

14. Rohatgi A. WebPlotDigitizer, Version 4.3. https://automeris.io/WebPlotDigitizer. Published 2020. Accessed 9/28/20.

15. Anderson BJ, McKee AD, Holford NH. Size, myths and the clinical pharmacokinetics of analgesia in paediatric patients. Clin Pharmacokinet. 1997;33(5):313–327.

16. Mingeot-Leclercq MP, Tulkens PM. Aminoglycosides: nephrotoxicity. Antimicrob Agents Chemother. 1999;43(5):1003–1012.

17. Riethmueller J, Ballmann M, Schroeter TW, et al. Tobramycin once-vs thrice-daily for elective intravenous antipseudomonal therapy in pediatric cystic fibrosis patients. Infection. 2009;37(5):424–431.

18. Landmesser KB, Autry EB, Gardner BM, Bosko KA, Schadler A, Kuhn RJ. Comparison of the predictive value of area under the curve versus maximum serum concentration of intravenous tobramycin in cystic fibrosis patients treated for an acute pulmonary exacerbation. Pediatr Pulmonol. 2021;56(10):3209–3216.

19. Akcan-Arikan A, Zappitelli M, Loftis LL, Washburn KK, Jefferson LS, Goldstein SL. Modified RIFLE criteria in critically ill children with acute kidney injury. Kidney Int. 2007;71(10):1028–1035.

20. Downes KJ, Rao MB, Kahill L, Nguyen H, Clancy JP, Goldstein SL. Daily serum creatinine monitoring promotes earlier detection of acute kidney injury in children and adolescents with cystic fibrosis. J Cyst Fibros. 2014;13(4):435–441.

21. Massie J, Cranswick N. Pharmacokinetic profile of once daily intravenous tobramycin in children with cystic fibrosis. J Paediatr Child Health. 2006;42(10):601–605.

22. Hennig S, Norris R, Kirkpatrick CM. Target concentration intervention is needed for tobramycin dosing in paediatric patients with cystic fibrosis--a population pharmacokinetic study. Br J Clin Pharmacol. 2008;65(4):502–510.

23. Touw DJ, Knox AJ, Smyth A. Population pharmacokinetics of tobramycin administered thrice daily and once daily in children and adults with cystic fibrosis. J Cyst Fibros. 2007;6(5):327–333.

